# Oligosymptomatic long-term carriers of SARS-CoV-2 display impaired innate resistance and high Spike-specific neutralizing antibodies

**DOI:** 10.1101/2022.11.19.22282546

**Authors:** Elena Montes-Cobos, Victoria C Bastos, Clarice Monteiro, João CR de Freitas, Heiny DP Fernandes, Clarice S Constancio, Danielle AS Rodrigues, Andreza MDS Gama, Vinicius M Vidal, Leticia S Alves, Laura Zalcberg-Renault, Guilherme S de Lira, Victor A Ota, Carolina Caloba, Luciana Conde, Isabela C Leitão, Amilcar Tanuri, Orlando DC Ferreira, Renata M Pereira, André M Vale, Terezinha M Castiñeiras, Dominique Kaiserlian, Juliana Echevarria-Lima, Marcelo T Bozza

**Affiliations:** Laboratório de Inflamação e Imunidade, Instituto de Microbiologia Paulo de Góes, Universidade Federal do Rio de Janeiro, Rio de Janeiro, Brazil; Laboratório de Imunologia Básica e Aplicada, Instituto de Microbiologia Paulo de Góes, Universidade Federal do Rio de Janeiro, Rio de Janeiro, Brazil; Laboratório de Biologia de Linfócitos, Instituto de Biofísica Carlos Chagas Filho, Universidade Federal do Rio de Janeiro, Rio de Janeiro, Brazil; Departamento de Doenças Infecciosas e Parasitárias, Faculdade de Medicina, Universidade Federal do Rio de Janeiro, Rio de Janeiro, Brazil; Laboratório de Virologia Molecular, Instituto de Biologia, Universidade Federal do Rio de Janeiro, Rio de Janeiro, Brazil; Laboratório de Imunologia Molecular, Instituto de Microbiologia Paulo de Góes, Universidade Federal do Rio de Janeiro, Rio de Janeiro, Brazil; Mucosal Immunity Vaccination & Biotherapies Laboratory, INSERM U1060, Université Claude Bernard Lyon 1, Centre hospitalier Lyon-Sud, Pierre-Benite, France

**Keywords:** SARS-CoV-2, host resistance, disease tolerance, persistent infection, IFN-I, Type 1/3 immunity, neutralizing antibodies

## Abstract

The vast spectrum of clinical features of COVID-19 keeps challenging scientists and clinicians. Control of pathogen load (host resistance) and prevention of tissue damage (disease tolerance) are essential for the outcome of infectious diseases. Both low resistance and high disease tolerance might result in long-term viral persistence, but the underlying mechanisms remain unclear. Here, we studied the immune response of immunocompetent COVID-19 patients with prolonged SARS-CoV-2 infection by immunophenotyping, cytokine and serological analysis. Despite viral loads and symptoms comparable to regular mildly-symptomatic patients, long-term carriers displayed weaker systemic IFN-I responses and fewer circulating pDCs and NK cells at disease onset. Type 1 cytokines remained low, while type-3 cytokines were in turn enhanced. Interestingly, the plasma of these patients showed a higher spike-specific neutralization capacity. The identification of very early distinct immune responses in long-term carriers adds up to our understanding on essential host protective mechanisms to ensure tissue damage control despite prolonged viral infection.

## INTRODUCTION

Patients infected with severe acute respiratory syndrome coronavirus 2 (SARS-CoV-2) have clinical presentations ranging from asymptomatic-mildly symptomatic (70-90%) to severe and critical (10-30%) ^1–4^. These different clinical outcomes, including the risk of COVID-19-related death, have been associated with age, gender, and underlying comorbidities, such as obesity and diabetes ^2,5,6^. Hospitalized patients with severe COVID-19 present an increase in inflammatory cytokines, monocytes and neutrophils, and a marked decrease in lymphocytes compared to patients with mild disease ^7–9^. Moreover, a significant fraction of patients with life-threatening COVID-19 present defects in type I IFNs (IFN-I) due to inborn mutations and auto-antibodies, pointing to a critical role of IFN-I in the immune response against SARS-CoV-2 ^10–12^. These distinct immune and inflammatory signatures are observed early after COVID-19 diagnosis, correlate with divergent disease trajectories and might have prognostic value ^9,13,14^.

The interplay of resistance to limit pathogen burden and maintenance of tissue homeostasis, also known as disease tolerance, is central to host and pathogen interactions and determines disease progression and outcome ^15,16^. Whilst resistance represents the ability to contain pathogen replication and spread in the host, disease tolerance reflects the capacity to limit deleterious effects of infectious agents and the associated inflammatory response. Indeed, critically ill COVID-19 patients present local and systemic inflammation with severe tissue dysfunction regardless of pathogen loads ^9,13,14^. Alternatively, immunosuppressed individuals, who exemplify the paradigm of low host resistance, display a variety of clinical presentations, from asymptomatic to severe ^17– 20^. These observations suggest that disease tolerance mechanisms controlling tissue homeostasis might underly a benign clinical course of SARS-CoV-2 infection and determine the fitness of COVID-19 patients ^21^.

Low resistance might impact SARS-CoV-2 clearance in multiple ways, leading to high viral titers in the upper-respiratory tract (URT), dissemination to other tissues, especially the lungs, or long-term virus persistence. Although viral persistence has been more frequently described in immunosuppressed patients, persistent URT infection and long-term virus shedding have been documented in immunocompetent patients with asymptomatic or mild COVID-19 as well ^22–26^. Most long-term carriers remained SARS-CoV-2 positive by qRT-PCR despite seroconversion, reinforcing the risk of continuous SARS-CoV-2 transmission ^23,27,28^. Defects in antigen-specific cytotoxic T cell responses were found in such patients ^29^, but the immune dynamics during the first days of infection remains unclear. Thus, the aim of this study is to gain insights into the immune mechanisms associated to prolonged SARS-CoV-2 infection in oligosymptomatic, immunocompetent subjects. First, we identified early pathophysiological immune markers that may predict COVID-19 progression. Overall, our study reveals alternative immune strategies to cope with SARS-CoV-2 infection, shedding light on the mechanisms of resistance and disease tolerance in COVID-19.

## RESULTS

### Demographic characterization of study cohort

Our study cohort is composed of individuals tested for SARS-CoV-2 infection at the Diagnostic Screening Center for COVID-19 of the Federal University of Rio de Janeiro (CTD-UFRJ) from April to December 2020. Weekly follow-up was offered to those subjects who tested positive for the presence of SARS-CoV-2 RNA by quantitative PCR with reverse transcription (RT-qPCR) in nasopharyngeal swab samples, until SARS-CoV-2 RNA was no longer detectable.

Initial studies performed in the CTD-UFRJ cohort found a median of SARS-CoV-2 RT-qPCR positivity around three weeks after symptoms onset ^22^. Day 21 after symptom onset (DSSO) was thus used as a putative threshold time point of viral clearance from the URT. From those individuals who volunteered to longitudinal follow-up testing, we selected 33 patients with persistent SARS-CoV-2 infection (P), representing long-term carriers, defined by detectable SARS-CoV-2 RNA (i.e. Ct < 38) at ≥ 21 DSSO. Thirty-two SARS-CoV-2 infected patients with a negative RT-qPCR (i.e. Ct ≥ 38) at ≤ 21 DSSO were also included as COVID-19 control study group, thereafter referred to as non-persistent (NP), and 24 age-and gender-matched asymptomatic, non-infected individuals (confirmed by the absence of spike-specific IgM and IgG in the plasma) were selected as the reference group (NI) (Fig. 1A). The proportion of men and women was equally distributed among the study groups. The median age distribution was 36 (±9.67) years of age for the NI group, 36 (±11.05) for the those of the NP group and 38 (±12.13) for patients with persistent SARS-CoV-2 infection (P). COVID-19 patients included in this study were either oligosymptomatic, presenting mild symptoms including fever, headache, cough, sneeze, anosmia or myalgia, or asymptomatic at the time of enrollment. Despite prolonged URT infection, long term carriers displayed neither a significantly longer duration of symptoms (Fig. 1B) nor augmented disease severity markers in the plasma (Fig. 1C). None of the patients of our cohort developed severe COVID-19 requiring hospitalization or oxygen therapy. The most common comorbidities reported by the patients were hypertension (15.63% of NP versus 27.28% of P), hypothyroidism (6.25% NP versus 12.12% of P), and diabetes mellitus (6.25% of NP versus 3.03% of P) (Table 1).

**Figure 1.**
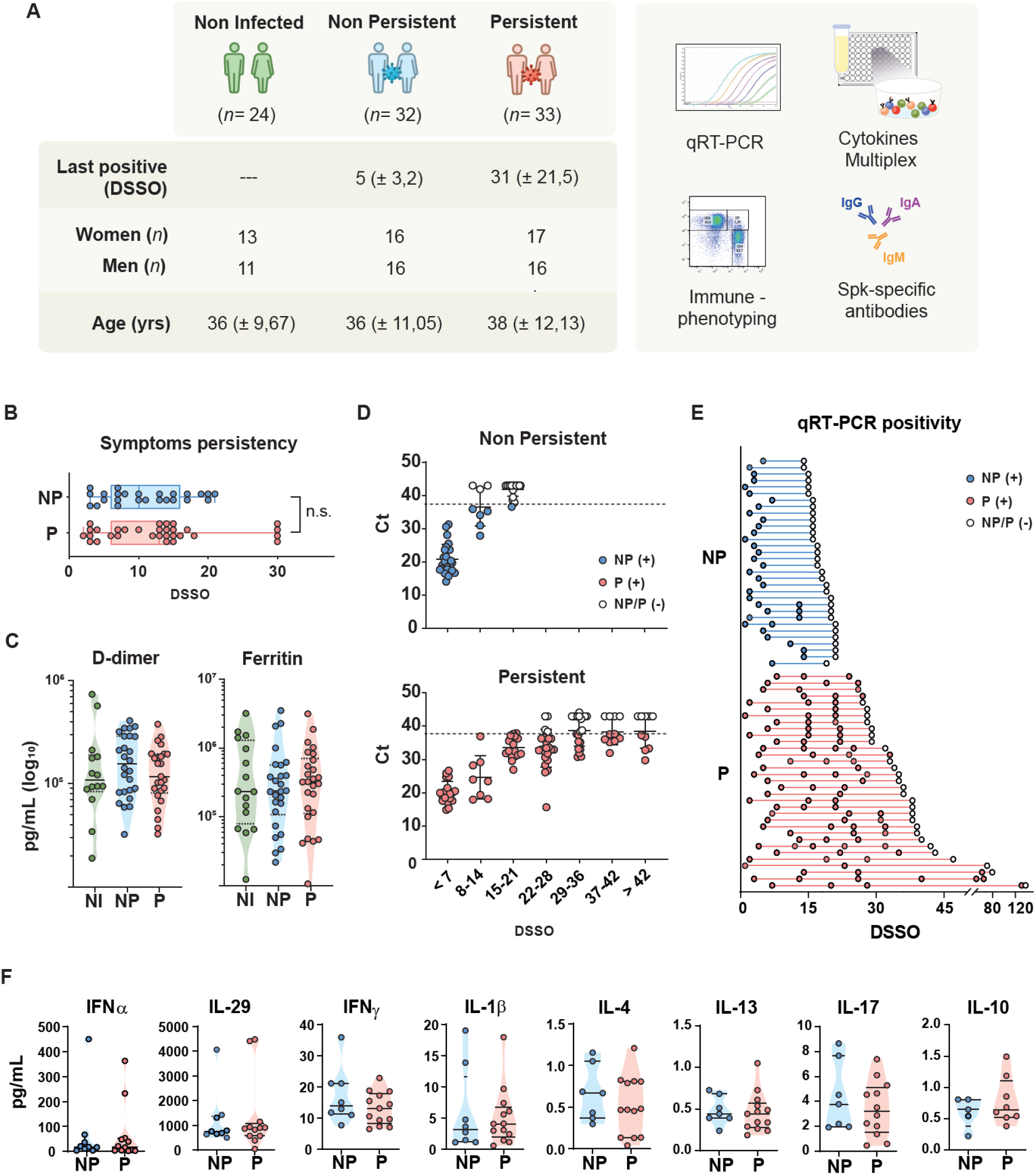
Overview of clinical features, viral loads, and mucosal immunity parameters of COVID-19 patients. (A) SARS-CoV-2 positivity by qRT-PCR in nasopharyngeal samples, sex and age of non-infected controls (NI), non-persistent (NP) and persistent (P) COVID-19 patients. Duration of symptoms of NP and P patients, represented as days after symptom onset (DSSO). Quantification of the plasma damage markers D-dimer and Ferritin in NI (*n*=14), NP (*n*=26), and P (*n*=24) at <10 DSSO by multiplex magnetics bead-based immunoassay. (D) Cycle threshold (Ct) values of qRT-PCR for SARS-CoV-2 target genes from nasopharyngeal samples from NP (*n*=32) and P (*n*=33) patients by DSSO. (E) SARS-CoV-2 positivity of NP (*n*=32) and P (*n*=33) patients by DSSO. (F) Quantification of IFNα, IL-29, IFNγ, IL-1b, IL-4, IL-13, IL-17, and IL-10 from URT samples in NP (*n*=9) and P (*n*=13) at <10 DSSO by multiplex immunoassay. Each dot represents a subject. Filled dots represent a positive qRT-PCR of nasopharyngeal samples for SARS-CoV-2, while empty dots represent a negative qRT-PCR. DSSO, Days since symptom onset.

**Table 1.** Demographics of patient cohort.

|  | Non persistent (n=32) | Persistent (n=33) | P value |
| --- | --- | --- | --- |
| ***Characteristics*** |  |  |  |
| Age, median (years) | 36 (±11.05) | 38 (±12.13) | 0.4606 |
| Sex, female | 16/32 (50%) | 17/33 (51.52%) |  |
| Last positive, median (DSSO) | 5 (±3.2) | 31 (±21.5) | <0.0001 |
| ***Comorbidities*** |  |  |  |
| Hypertension | 5/32 (15.63%) | 9/33 (27.28%) |  |
| Diabetes Mellitus | 2/32 (6.25%) | 1/33 (3.03%) |  |
| Hypothyroidism | 2/32 (6.25%) | 4/33 (12.12%) |  |
| Others | 4/32 (12.50%) | 5/33 (15.16%) |  |

### SARS-CoV-2 persistency does not depend on viral load or mucosal cytokines

As SARS-CoV-2 enters the organism via the URT, early local immunity at the nasopharyngeal mucosa may be important for fast and efficient viral clearance. Therefore, we first analyzed early viral loads and immune parameters in the nasopharyngeal mucosa of patients that eventually developed prolonged SARS-CoV-2 infection. Viral titers, determined by qRT-PCR, in nasopharyngeal swabs of long-term carriers were similar to those of non-persistent COVID-19 patients during the first days of infection (≤ 7 DSSO). Long-term carriers presented detectable viral RNA in the URT for up to 4 weeks, albeit with slightly higher Ct values (Fig. 1D). Strikingly, a group of patients still tested positive by RT-qPCR for longer than 8 weeks, with the latest positivity result being at 134 DSSO in one patient (Fig. 1E).

Next, we used a multiplex assay to compare a set of 47 immune mediators, including IFNα, IL29, IFNγ, IL-1β, IL-4, IL-13, IL-17A and IL-10, in nasopharyngeal swab samples collected at ≤ 10 DSSO from a group of non-persistent patients (NP, *n*=9) and long-term carriers (P, *n*=13). The results did not show any differences in early cytokines between both groups (Fig. 1F, Suppl. Fig. 1A). These results suggest that early alterations of these mucosal cytokines at the entry site of SARS-CoV-2 do not drive viral persistence.

### Long-term carriers present reduced systemic type I IFN responses

Early dysregulated systemic inflammatory and antiviral responses have been pointed out as potential drivers of distinct clinical progression of COVID-19 ^9,10,13^. To gain insights into specific immune mechanisms leading to prolonged SARS-CoV-2 infection, we analyzed innate immune cells and soluble mediators in non-infected individuals, non-persistent COVID-19 patients, and long-term carriers early after disease onset (≤ 10 DSSO). Long-term carriers displayed a different distribution pattern of circulating monocyte populations, characterized by a drop in the frequency of classical (lin^-^CD14^+^CD16^-^) monocytes and a modest increase in the percentage of non-classical (lin^-^CD14^-^CD16^+^) monocytes (Fig. 2A). Additionally, we found a significant reduction of pDC frequencies (lin^-^CD14^-^CD304^+^) in the blood of these patients (Fig. 2B), which may be due to lower numbers of circulating pDCs or to higher tissue recruitment. Nonetheless, we detected augmented expression of HLA-DR on this cell population, indicative of a more mature phenotype and a predisposition to apoptosis ^30^. Other monocyte and dendritic cell populations did not vary between both groups of patients (Suppl. Fig. 3A).

**Figure 2.**
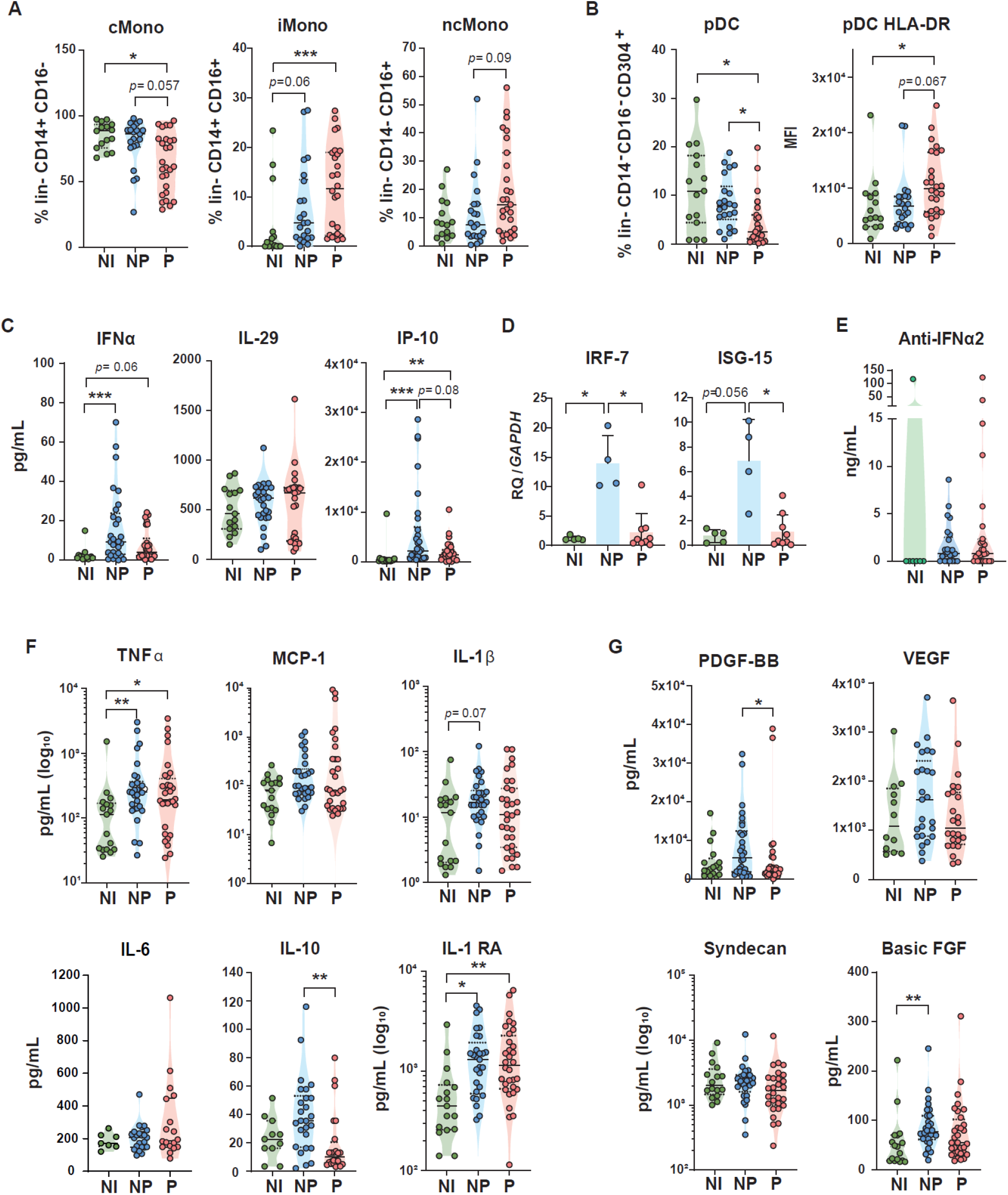
Early innate immune profiles distinguish between persistent and non-persistent patients. Immunophenotyping of innate immune cell populations in PBMC from NI (*n*=15), NP (*n*=22) and P (*n*=27) at <10 DSSO, depicting (A) LIVE/DEAD^+^CD3^-^CD19^-^CD56^-^ (lin^-^) CD14^+^CD16^-^ classical monocytes (cMono), CD14^+^CD16^+^ intermediate monocytes (iMono), CD14^-^CD16^+^ non-classical monocytes (ncMono), (B) lin^-^CD14^-^CD16^-^CD304^+^ plasmacytoid cells (pDC) and activated pDCs (MFI of HLA-DR in pDC). Quantification of plasma cytokines in NI (*n*=17), NP (*n*=27) and P (*n*=32) at <10 DSSO by multiplex magnetics bead-based immunoassay. Expression of IRF-7 and ISG-15 in PBMC of NP and P at < 10 DSSO by PCR. Each dot represents a different subject. Statistical significance was calculated using Mann-Whitney test or Kruskal-Wallis analysis followed by Dunn post-test and indicated by *p % 0.05; **p % 0.01; and ***p % 0.001. DSSO, Days since symptom onset.

Since pDCs are the first and major IFN-I producers during viral infections ^31,32^, it was not unexpected that plasma concentrations of IFNα and IP-10 (CXCL10) in long-term carriers did not increase as much as in non-persistent patients (Fig. 2C). Expression analysis of the interferon-inducible genes (ISGs) IRF7 and ISG-15 in PBMCs also showed reduced induction of both genes in patients from the P group compared to those of the NP group (Fig. 2D). No differences were observed in the plasma concentrations of IFNλ (IL-29) (Fig. 2C), IFNβ or IFNω (Suppl. Fig. 3B), nor in the occurrence of anti-IFNα2 autoantibodies (Fig. 2 E). Neither did we find changes in most of the inflammatory mediators predictive of poor outcome in COVID-19, such as TNFα, MCP-1 (CCL2), IL1β, IL-6 or IL-1 RA (Fig. 2F, Suppl. Fig. 3B) ^7–9^. Only plasma concentrations of the regulatory cytokine IL-10 were lower in long-term carriers than in non-persistent patients.

Furthermore, we titrated plasma levels of molecules involved in tissue damage and repair at disease onset, some of which have been previously connected to disease tolerance ^33^. In contrast to non-persistent patients, long-term carriers showed no increase in tissue growth factors like PDGF-BB, basic FGF and VEGF, indicating that systemic tissue repair mechanisms were not induced in these patients (Fig. 2F). Syndecan-1, involved in modulation of inflammation and tissue repair and a hallmark of endothelial damage ^34^, was comparable in all groups. Collectively, our results show altered innate immune responses, a weaker antiviral state and limited induction of tissue repair during the first days of prolonged SARS-CoV-2 infection.

### Type 1 responses shift to type 3 immunity in long-term carriers

The immune system orchestrates distinct resistance mechanisms depending on the nature of the infectious agent, the site of infection and the time after infection onset. Type 1 immunity, mediated by IFNγ, NK cells, T helper 1 (Th1) lymphocytes and cytotoxic T cells, is primarily induced in response to intracellular pathogens, such as viruses. We thus analyzed type 1 immune responses in our cohort of long-term carriers. First, we detected lower frequencies of circulating NK cells in persistently-infected COVID-19 patients compared to non-persistent ones (Fig. 3A). We also observed that the proportion of circulating naïve CD4^+^ was slightly reduced in patients of the P group (Fig. 3B). These immunophenotypic alterations were in line with a drop in plasmatic IL-7 (Fig. 3C), as this cytokine contributes to NK cell and naïve/memory T cell development and survival ^35^. Additionally, we stimulated total PBMCs *in vitro* with anti-CD3/CD28 beads in order to analyze the inducible cytokine production by circulating CD4^+^ and CD8^+^ T cells. Polyclonally-activated PBMCs from long-term carriers showed elevated amounts of TNFα produced by T helper cells and Granzyme B by cytotoxic T cells, while the synthesis of these cytokines in the non-persistent COVID-19 patients was not different from that of non-infected individuals at this early time point (≤ 10 DSSO) (Fig. 3D). Further analysis of systemic hallmarks of type 1 immunity revealed significantly lower IFNγ and IL-12 concentrations in the plasma of long-term carriers (Fig. 3E), but higher circulating levels of IL-15, a broadly pleiotropic cytokine with stimulating effects on NK cells and T lymphocytes.

**Figure 3.**
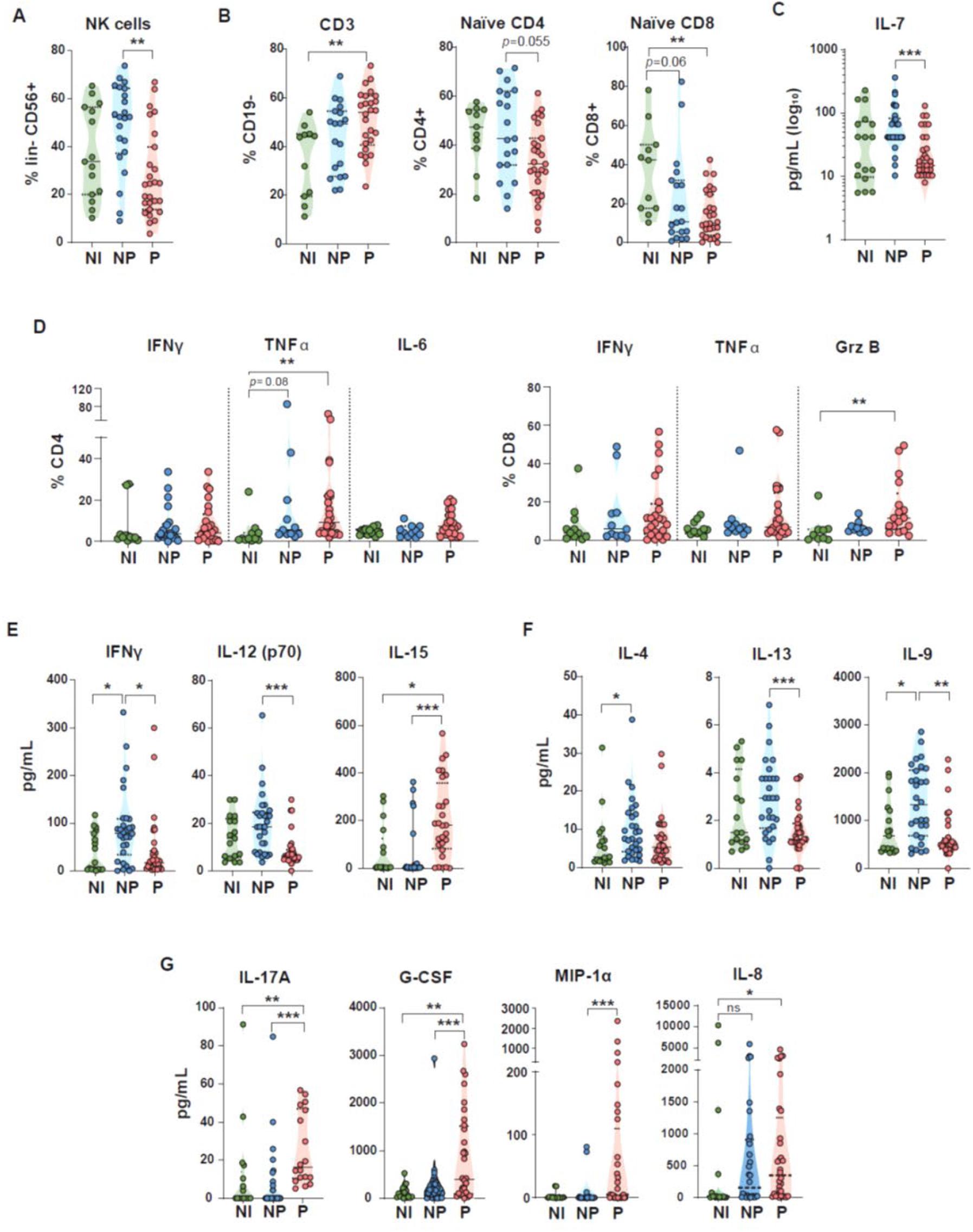
Dampened type 1 immunity in persistent patients. Immunophenotyping of immune cell populations in PBMC from NI (*n*=15), NP (*n*=22) and P (*n*=27) at <10 DSSO depicting (A) CD3^-^CD56^+^ natural killer cells (NK cells), (B) CD19^-^CD3^+^ T lymphocytes (CD3), CD4^+^CD27^+^CD45RA^+^ naive CD4 T lymphocytes (Naive CD4), and CD8^+^CD27^+^CD45RA^+^ naive CD8 T lymphocytes (Naive CD8). (C) Quantification of plasma IL-7 in (*n*=17), NP (*n*=27) and P (*n*=32) at <10 DSSO by multiplex magnetics bead-based immunoassay. (D) Frequencies of CD4^+^ T lymphocytes producing IFNγ, TNFα and IL-6 and CD8^+^ T lymphocytes producing IFNγ, TNFα and Granzyme B (Grz B) after polyclonal in vitro stimulation with anti-CD3/CD28 beads of PBMCs from (*n*=12), NP (*n*=11) and P (*n*=23). Quantification of plasma (E) IFNγ, IL-12, IL-15, (F) IL-4, IL-13, IL-9, (G) IL-17A, G-CSF, MIP-1α, and IL-8 in (*n*=17), NP (*n*=27) and P (*n*=32) at <10 DSSO by multiplex magnetic bead-based immunoassay. Statistical significance was calculated using Mann-Whitney test or Kruskal-Wallis analysis followed by Dunn post-test and indicated by *p % 0.05; **p % 0.01; and ***p % 0.001. DSSO, Days since symptom onset.

Considering the reduced type 1 immunity taking place only in the of patients subsequently evolving towards a persistent infection, we next explored whether long-term carriers presented a functional deviation to type 2 or type 3 immunity. Analysis of plasma cytokines showed that, while type 2-associated cytokines, such as IL-4, IL-13 and IL-9, were poorly represented in the plasma of patients with prolonged infection, the concentration of IL-17A was strongly increased (Fig. 3F, G). Soluble mediators responsible for neutrophil development and recruitment, such as G-CSF, MIP-1α (CCL4) and IL-8 (CXCL8) were also augmented, indicating a more prominent type 3 immune profile. These observations suggest a shift from type 1 immunity to type 3 immune responses in patients with persistent SARS-CoV-2 infection.

### Patients developing persistent infection display an early distinct immune signature

Unsupervised cluster analysis of plasma cytokines at ≤ 10 DSSO revealed at least two groups of immune mediators that were differentially regulated in patients with normal resolution of infection compared to those with long-term infection (Fig. 4A). Cluster 1, composed of IL-6, IL-8, IL-17A, G-CSF, IL-15 and MIP-1α, was overrepresented in most long-term carriers, whereas the cytokines from cluster 5, containing IL-9, IL-10, IFNγ and IL-12, were reduced. Additionally, we combined plasma cytokine data with immunophenotyping data from the same time points (Fig 4B and Suppl. Fig.4A, B). The correlation matrix of soluble proteins and immune cell subtypes revealed a positive correlation of effector CD8^+^ T cells with Th1 and Th2 cytokines, which, in turn, inversely correlated with the activation of alternative monocytes and pDCs (Fig. 4B). Furthermore, unsupervised cluster analysis on cytokine data by t-Distributed Stochastic Neighbor Embedding (t-SNE) identified four different groups of patients (Suppl. Fig. 4B). All non-infected controls gathered in one cluster, whereas long-term carriers were distributed in three different groups along with non-persistent patients, irrespective of age, gender or comorbidities. Finally, fold change importance analysis performed on NP versus P patients highlighted IL-17A, MIP1α, IL-15, IL-8 and IFNγ as the top five cytokines defining SARS-CoV-2 persistency (Fig. 4C). Altogether, bioinformatic analysis strongly suggests that a combination of early blood markers could predict prolonged SARS-CoV-2 infection.

**Figure 4.**
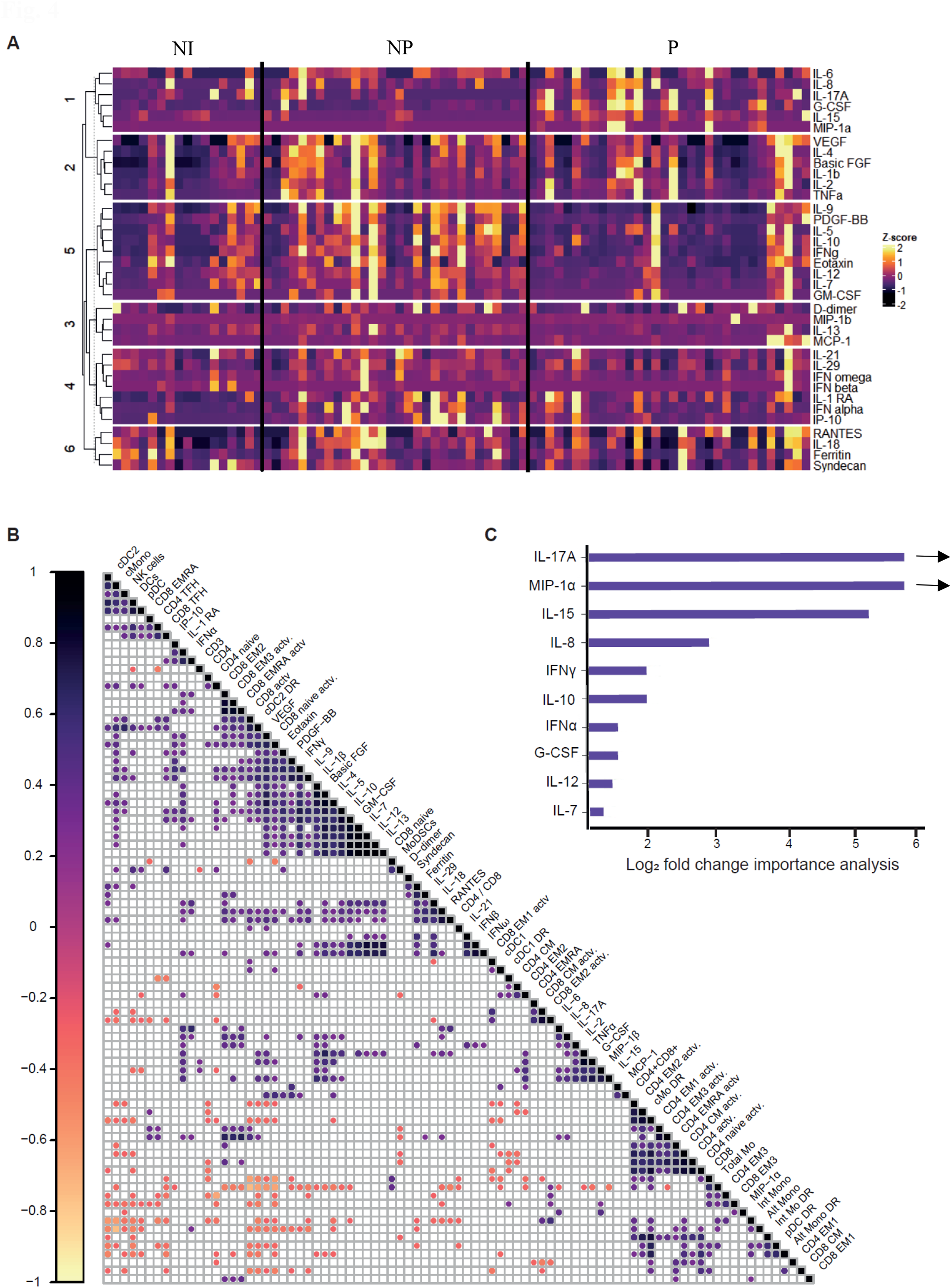
Early immune signature of persistent patients. (A) Heat map of cytokine concentration in serum from NI, NP and P at <10 DSSO measured by multiplex assay. K-means clustering was used to determine cytokine clusters 1-6 (cluster 1, n = 6; cluster 2, n = 6; cluster 3, n = 4; cluster 4, n = 7; cluster 5, n = 9; cluster 6, n = 4). (B) Correlation matrix across PBMC immunophenotyping and cytokines concentrations from NI, NP and P at <10 DSSO. Only significant correlations (p<0.05) are represented as dots. Pearson’s correlation coefficients from comparisons of cytokine measurements within the same patients are visualized by color intensity. (C) Fold Change importance analysis between NP and P, calculated using the gtools package in R. IL-17A and MIP-1α have median equals zero in NP, resulting in an infinite fold change, which is represented by the arrow.

### Longitudinal immune profiling of patients with prolonged SARS-CoV-2 infection

Next, we analyzed the cytokine dynamics over the course of infection in the patients of our cohort (Fig. 5A, Suppl. Fig. A). Both IFNα and its target chemokine IP-10 progressively decreased over time in long-term carriers. Suppression of IP-10 has been proposed to be a mechanism of pathogen evasion and persistence in different infection models ^36^. This trend could also be observed for IFNγ, thus ruling out the possibility of a boost of systemic IFN responses leading to final viral clearance from the URT. Other cytokines went through a transient increase during the second week after symptom onset and finally dropped by the end of infection. This effect was particularly pronounced for TNFα, IL-12, IL-17A and IL-8, suggesting that these cytokines might contribute to SARS-CoV-2 clearance in the absence of efficient type I IFN responses.

**Figure 5.**
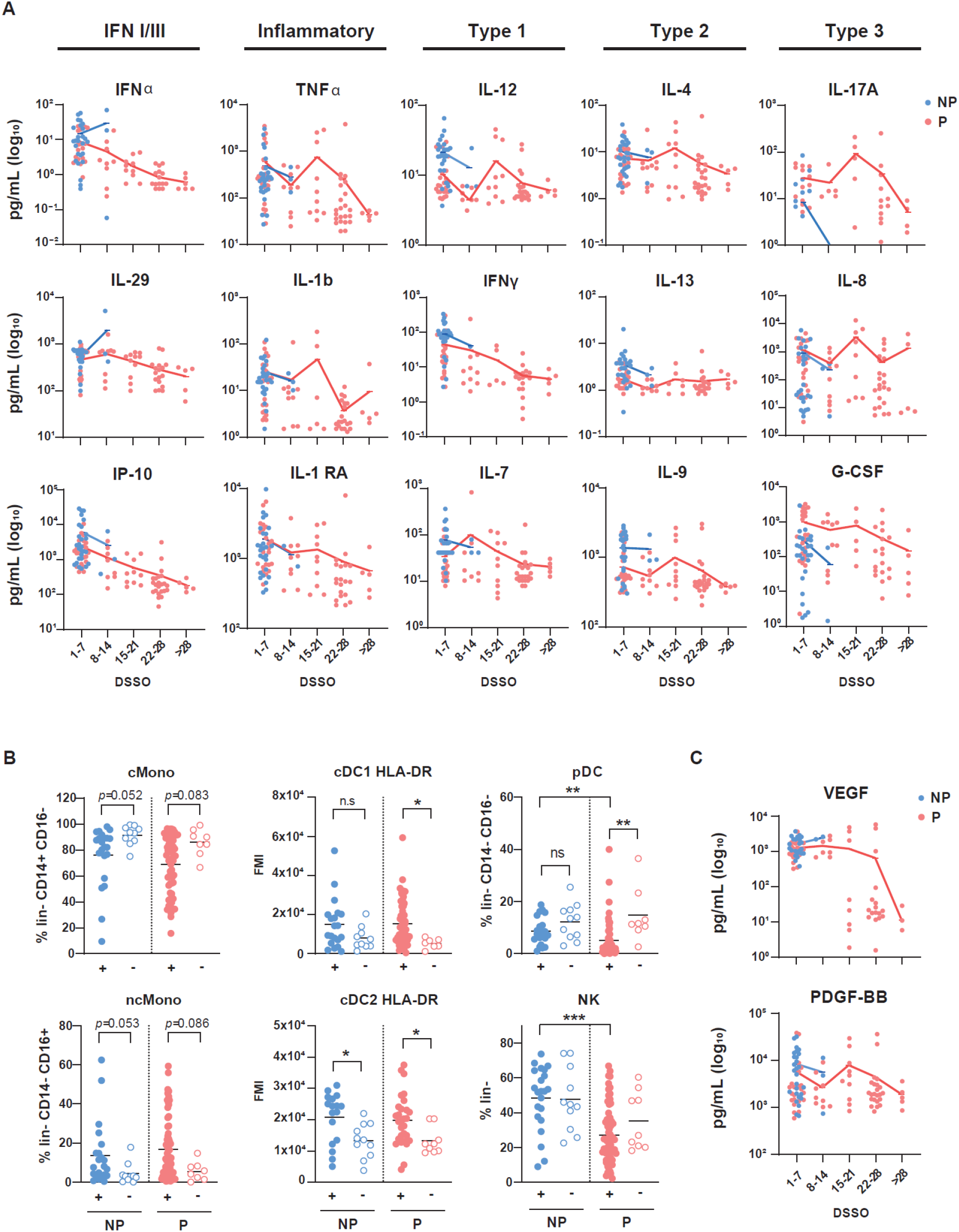
Longitudinal immune profiling of persistent patients. (A) Weekly longitudinal quantification of plasma cytokines in non-persistent (NP, blue) and persistent (P, red) patients by multiplex immunoassay. (B) Longitudinal immunophenotyping of PBMCs from NP and P depicting frequencies of CD14^+^CD16^-^ classical monocytes (cMono) and CD14^-^CD16^+^ non-classical monocytes (ncMono), mean fluorescence intensity (MFI) of HLA-DR in CD14^-^CD16^-^ CD11c^+^CD141^+^ (cDC1) and CD14^-^CD16^-^CD11c^+^CD1c^+^ (cDC2), and frequencies of CD14^-^CD16^-^ CD304^+^ plasmacytoid cells (pDC) and CD3-CD56^+^ natural killer cells (NK). Filled dots represent individual samples longitudinally collected at different time points until resolution of infection from patients positive by qRT-PCR for SARS-CoV-2, while empty dots represent samples from convalescent patients coinciding with the first negative qRT-PCR for SARS-CoV-2. (C) Weekly longitudinal quantification of VEGF and PDGF-BB in NP and P patients by multiplex magnetics bead-based immunoassay. Statistical significance was calculated using Mann-Whitney test or Kruskal-Wallis analysis followed by Dunn post-test and indicated by *p % 0.05; **p % 0.01; and ***p % 0.001. DSSO, Days since symptom onset.

To visualize the interplay between these cytokines and viral loads, we depicted cytokine concentrations along with Ct values (represented as Ct-^1^) for the eight individual patients with more than three longitudinal samples (Suppl. Fig. 6A). Instead of a unique immune profile, we found distinct cytokine and Ct dynamics. For instance, patient P11 had almost absent type 1 immune responses and elevated concentrations of inflammatory and type 3 cytokines at the beginning of infection, and only IL-1 RA increased at later time points. Alternatively, patient P8 began with overall lower cytokine concentrations that progressively increased, peaking at the third week after disease onset and shortly before viral clearance. Another illustrative example was patient P24, who remained infected by SARS-CoV-2 for the longest period of time (134 DSSO) and displayed strong viral replication during the first two weeks of infection, without evident alterations in plasma cytokine dynamics. Later on, an increase in TNFα, IFNγ and IL-8 was apparent, coinciding with the stabilization of viral titers and final viral clearance. These data suggest that each long-term infected individual copes with viral persistence in a different manner. Furthermore, we studied the behavior of different immune populations longitudinally.

Generally, the immunophenotypic changes found during the first 10 days of SARS-CoV-2 infection were maintained until the virus was cleared from the nasopharynx (Fig. 5B, Suppl. Fig. 7A). Frequencies of classical, intermediate and non-classical monocytes only returned to control levels at time points where the virus was no longer detectable in the URT, and a similar trend was found for dendritic cell activation, monitored by the surface expression of HLA-DR. Additionally, most long-term carriers displayed lower pDC and NK cell frequencies all along the infection period.

Finally, in order to determine whether prolonged infection leads to progressive tissue damage, we quantified plasma concentrations of growth factors associated with wound repair mechanisms. While VEGF concentrations acutely dropped at the second week after symptom onset, some patients showed a transient increase in PDGF-BB and basic FGF around this time point and stabilized later on, following the same trend as most inflammatory mediators (Fig. 5C, Suppl. Fig. 5A). Altogether, these results suggest that prolonged SARS-CoV-2 infection with low viral loads does not lead to persistent inflammation or to systemic tissue damage.

### SARS-CoV-2 persistency favors the generation of Spike-specific neutralizing antibodies

Adequate innate immune activation is essential for the development of antigen-specific adaptive immunity. Therefore, we next evaluated whether the distinct early immune profile that we found in long-term carriers would have consequences for the development of immunological memory. First, we analyzed different memory T cell populations over time. Some patients with persistent infection displayed higher frequencies of effector memory (EM) and terminally differentiated (EMRA) CD4^+^ T cells over the course of infection (Fig. 6A, Suppl. Fig. 7A). We also observed sustained elevated frequencies of circulating central memory (CM) and EM CD8^+^ T cells until virus remission in this group of patients. Furthermore, in order to see whether long-term carriers were able to develop SARS-CoV-2-specific effector T cells, we isolated PBMCs at the convalescent phase and re-stimulated them *in vitro* with a pool of peptides spanning the Spike protein of the alpha variant of SARS-CoV-2. The results show no significant differences in actively proliferating T cells (Ki67^+^) between control COVID-19 patients and long-term carriers. Cytokine release by CD4^+^ and CD8^+^ T cells was comparable as well, except from IL-10 production by CD4^+^ CD25^+^ T cells, which was once again lower in the persistent group (Fig. 6B, C). These results indicate that persistent infection does not interfere with the development of antigen-specific T cell-mediated immune memory, and suggest that protective cellular mechanisms against re-infection are not impaired in long-term carriers.

**Figure 6.**
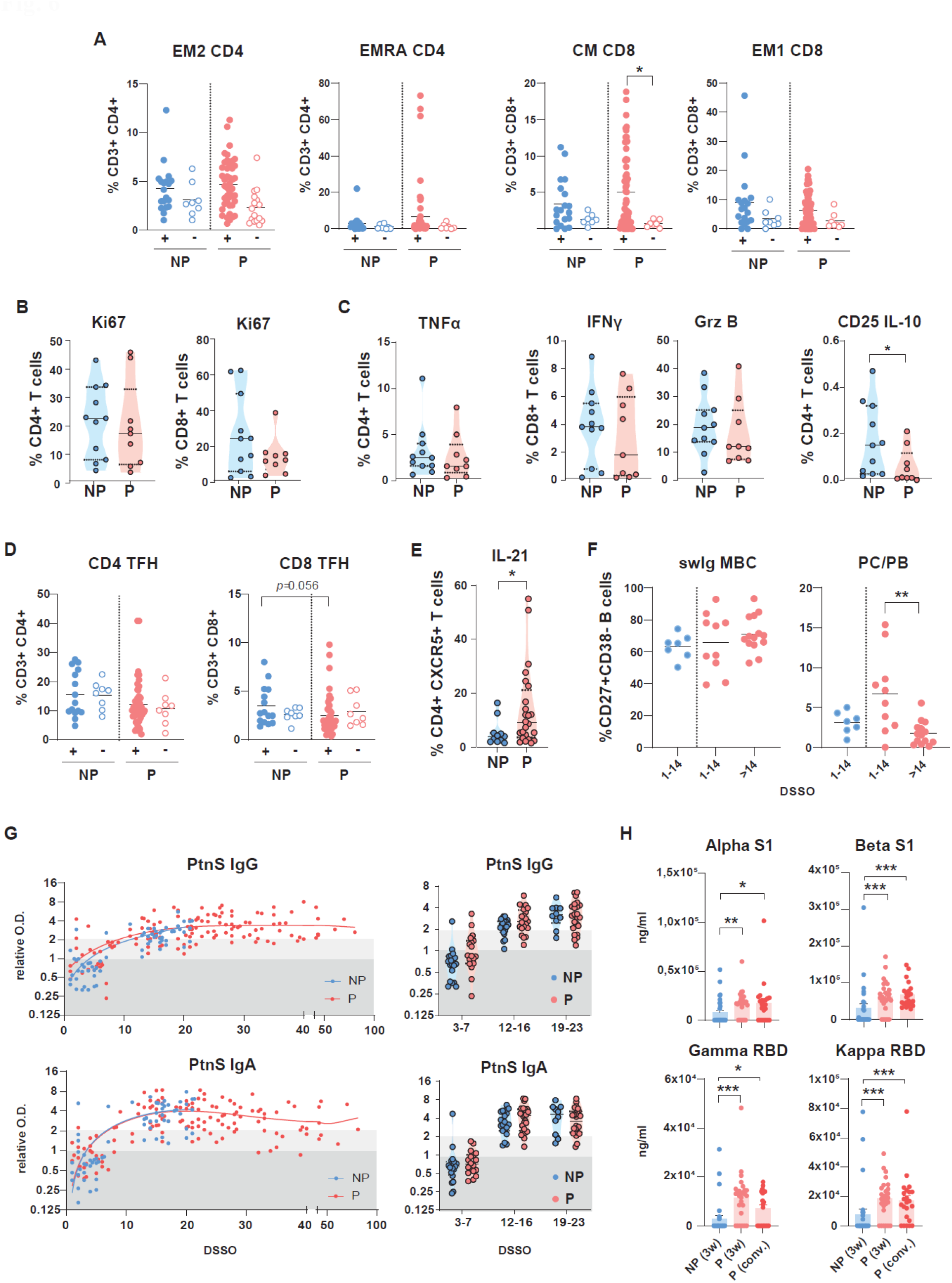
Profiling of cellular and serological adaptive response of persistent patients. (A) Longitudinal immunophenotyping of PBMC from NP and P depicting CD4^+^CD27^-^CD45RA^-^ CCR7^+^ type 2 effector memory T cells (EM2), CD4^+^CD27^-^CD45RA^+^CCR7^-^ terminally differentiated effector memory T cells (EMRA), CD8^+^CD27^+^CD45RA^-^CCR7^+^ central memory T cells (CM), and CD8^+^CD27^-^CD45RA^-^CCR7^-^ type 1 effector memory T cells (EM1). Filled dots represent individual samples longitudinally collected until resolution of infection from NP and P patients positive by qRT-PCR for SARS-CoV-2. Empty dots represent samples from convalescent patients coinciding with the first negative qRT-PCR for SARS-CoV-2. (B, C) Immunophenotyping of SARS-CoV-2 reactive CD4 and CD8 T cells after *in vitro* stimulation of PBMC from NP (*n*=11) and P (*n*=9) after viral clearance with peptides spanning the Spike protein of the alpha variant of SARS-CoV-2. Percentage of (B) CD4+ and CD8^+^ cells expressing Ki67, (C) CD4^+^ cells expressing TNFα, CD8^+^ cells expressing IFNγ and Granzyme B (Grz B), and CD4^+^CD25^+^ cells expressing IL-10. (D) Longitudinal immunophenotyping of PBMC from NP and P depicting CD3^+^CD4^+^CXCR5^+^ and CD3^+^CD8^+^CXCR5^+^ follicular T (TFH) cells. (E) Percentage of CD4^+^CXCR5^+^ T cells expressing IL-21 after polyclonal *in vitro* stimulation of PBMC from NP (*n*=11) and P (*n*=24) at <14 DSSO with anti CD3/CD28 beads. (F) Longitudinal immunophenotyping of PBMC from NP (*n*=7) and P (*n*=10) depicting CD27^+^CD38^-^ switched immunoglobulin (swIg) memory B cells (MBC) and plasmablast/plasma cells (PB/PC). (G) Longitudinal assessment of Spike-specific IgG and IgA antibodies in plasma from P and NP by ELISA assay. (H) Titers of neutralizing antibodies against Alpha S1, Beta S1, Gamma RBD, and Kappa RBD variants of SARS-CoV-2 in plasma from NP and P around 21 DSSO and P by the time of viral clearance by multiplex neutralization assay. Statistical significance was calculated using Mann-Whitney test or Kruskal-Wallis analysis followed by Dunn post-test and indicated by *p % 0.05; **p % 0.01; and ***p % 0.001. DSSO, Days since symptom onset.

Next, we aimed to study the development of antigen-specific humoral responses. We started analyzing follicular helper T cells (TFH), as they contribute to humoral immunity by delivering the necessary signals to B cells to enter germinal center and go through class-switch and affinity maturation. Although we did not find changes in the frequencies of circulating CD4^+^ TFH cells (Fig. 6D), polyclonal stimulation of PBMCs collected at ≤ 14 DSSO revealed that this T cell population produced higher amounts of IL-21 in long-term carriers, being this cytokine essential for B cell help (Fig. 6E). Furthermore, we compared the composition of the B cell compartment and the development of humoral responses in patients with normal or delayed resolution of infection. We did not observe significantly different frequencies of circulating mature, class-switched B cells, although the proportion of circulating plasmablasts/plasma cells dropped in long-term carriers after 14 DSSO (Fig. 6F, Suppl. Fig. 6A). Serological analysis did not identify significant alterations in the production of nucleocapsid (N) protein-specific IgG (Suppl. Fig. 8A) or spike protein-specific IgM, IgG and IgA antibodies over time (Fig. 6G, Suppl. Fig. 8A). However, plasma of long-term carriers displayed higher titers of circulating neutralizing antibodies against all analyzed SARS-CoV-2 variants three weeks after symptoms onset, and at least until the patients reached the convalescent phase (Fig. 6H, Suppl. Fig. 8B). These data suggest that development of systemic N protein-of spike protein-specific humoral responses do not seem to be sufficient for viral clearance during primary SARS-CoV-2 infection, although it may confer better protection against re-infection in long-term carriers.

## DISCUSSION

Our cohort comprises a group of oligosymptomatic patients with persistent infection who, by definition, have low resistance and high disease tolerance. Here we show that low resistance in these patients is likely due to impaired innate antiviral immunity, as no major defects in adaptive immunity were found. Furthermore, we observed divergent cytokine profiles systemically and in the nasopharyngeal mucosa, in line with other studies recently published ^37^. Major differences in cytokines, chemokines and growth factors were found in the plasma of patients with prolonged course of infection early after disease onset. However, although relevant mucosal cytokines, such as IL-17A and IL-10, were not altered in the nasopharynx, we cannot exclude the contribution of cytokines with only transient induction or other mucosal mediators not studied here.

Systemically, long-term carriers displayed decreased frequencies of circulating classical monocytes and pDCs shortly after symptom onset, being the latter essential for the control of viral replication through the rapid release of IFN-I ^31,32,38^. Early IFN-I triggers different mechanisms targeting virally infected cells, namely the expression of antiviral proteins, the activation of NK cells and the initiation of Th1 responses. Each one of these elements was underrepresented in those COVID-19 patients who later on presented persistent infection. Thus, our data suggest that early systemic immunological patterns may indicate future persistency of SARS-CoV-2 infection in immunocompetent patients.

Moreover, long-term carriers displayed an immunological shift characterized by a low systemic Th1/2 signature, increasing in turn IL-17A, IL-8 and other neutrophil-recruiting chemokines. In line with our data, mouse models of chronic infection by TMEV also link Th17 cells to viral persistence ^39^. Impaired IFN-I responses and an enhanced type 3 signature are traits that had been previously associated with severe COVID-19 ^9–11^. Conversely, inflammatory and Th1/2 responses are elevated in patients with severe symptoms but low in oligosymptomatic long-term carriers, which suggests their involvement in the pathogenesis of COVID-19 ^7–9,13^. Further studies to better characterize the correlation of these immunological patterns with infectious disease outcome are warranted.

The engagement of disease tolerance mechanisms, like enhanced tissue repair, metabolic adaptations or immune regulation, may also lead to distinct disease trajectories ^40,41^. Our data could not identify enhanced systemic tissue repair or regulatory responses during the course of prolonged SARS-CoV-2 infection. The immunoregulatory cytokine IL-10, which is commonly linked to immunological tolerance, but also a well-accepted disease marker in COVID-19 ^42,43^, was particularly low in long-term carriers. In order to understand the physiological meaning of this effect and the involvement of additional disease tolerance mechanisms, further analysis should be conducted.

In our study, long-term SARS-CoV-2 carriers had higher titers of neutralizing Spike-specific antibodies already three weeks after symptom onset. First, this indicates that systemic neutralizing antibodies are not sufficient for viral clearance from the URT during primary infection. Type I IFNs may contribute to this effect, since they have been associated with modulation of B cell responses and antibody production in the context of viral infection ^44,45^. Furthermore, patients with severe COVID-19, who may remain infected for extended time periods and present dysregulated IFN-I and type 3 responses, have also been shown to produce higher titers of neutralizing antibodies ^46^. Together with our data, this suggests that dampened IFN-I responses and/or strong type 3 immunity in persistent SARS-CoV-2 infection might confer advantages against re-infection. On the other hand, these antibodies might also contribute to viral persistence via antibody-dependent enhancement or modulate COVID-19 immunopathology via Fc-receptors. Hence, analysis of the functionality of the antibodies elicited during long-term infection, and longer follow-up of the adaptive cellular and humoral immunity in these patients could be valuable for COVID-19 treatment and vaccine design and development.

Given the out-care characteristic of our cohort and the techniques available, our investigation of viral RNA was restricted to nasopharyngeal samples. Assessment of viral titers in various tissues would allow for a better understanding of whether long-term carriers display systemic low resistance, or if this phenomenon is restricted to the mucosal site. Although we did not find increased damage and tissue repair markers in the blood or in the nasal mucosa, we cannot exclude that other SARS-CoV-2 target tissues, such as the lungs or the gut, have altered disease tolerance or support viral replication. To study disease tolerance and viral presence in such locations, animal models for SARS-CoV-2 would be helpful. Furthermore, our samples are limited and were collected in a specific timing and geographical context of the pandemic. Nonetheless, to our knowledge, the present study constitutes the characterization of immune parameters in the larger number of immunocompetent oligosymptomatic patients with persistent infection of SARS-CoV-2 so far. Still, sample size is critical when studying such a heterogeneous disease as COVID-19, and, therefore, validation of our data in larger, independent cohorts would be informative.

Collectively, our study provides a thorough analysis of the immune dynamics during viral persistency in COVID-19 patients. Over the past two years, important progress has been made in controlling the pandemic, but the vaccines available, though successful in limiting disease, are not sterilizing and do not completely prevent transmission of new SARS-CoV-2 variants ^47^. In COVID-19 as well as in other viral infections, oligosymptomatic and asymptomatic carriers represent the main vector of viral transmission. Therefore, long-term immunocompetent infected individuals are potential long-term spreaders and may, additionally, facilitate intra-host evolution of SARS-CoV-2 and other viruses ^17,22,23^. This reinforces the need for a better understanding of the immune mechanisms of viral control in these patients. All in all, our study identifies a set of early plasma markers associated with prolonged infection and reveals alternative immunological strategies to deal with viral infection without major tissue damage.

## Data Availability

All data produced in the present study are available upon reasonable request to the authors

## Acknowledgments

This work was partially supported by ANRS | Maladies infectieuses émergentes/INSERM grant (MUCOVID-007) (to DK and MTB); by CAPES, Edital de Seleção Emergencial II CAPES grant 88887.507381/2020-00 (to MTB); by Fundação de Amparo à Pesquisa do Estado do Rio de Janeiro (FAPERJ), grant E-26/201.128/2022 (272688) and E-26/211.564/2019 (252360) (to MTB), E-26/010.002434/2019 and E-26/210.178/2020 (to AT), E-26/203.002/208 (to A.M.V.); by Conselho Nacional de Desenvolvimento Científico e Tecnológico (CNPq) grant 312477/2021-0 (to MTB) and grants 439649/2018-8 and 316796/2021-2 (to A.M.V.); Instituto Serrapilheira (to AT).

EM-C; VCB; GSL; VAO were supported by fellowships from CAPES (88887.507381/2020-00).

CM; CC; LC were supported by fellowships from FAPERJ

DASR was supported by a fellowship from CNPq (DTI-A; 401209/2020-2).

ICL was supported by a fellowship from CNPq

JCRF and LZR were supported by fellowships from ANRS/INSERM (MUCOVID-007)

HDRF; CSC; VMV; AMS were supported by fellowships from CAPES

## Author contributions

Conceptualization: EM-C; VCB; CM; AT; DK; AMV; TMC; JE-L; MTB Methodology: EM-C; VCB; CM; JE-L; MTB

Investigation: EM-C; VCB; CM; JCRF; HDRF; CSC; DASR; AMS; VMV; LSA; LZR; GSL; VAO; CC; LC; ICL; JE-L

Formal analysis: EM-C; VCB; CM; JCRF; CC; DASR; RMP; AMV; JE-L; MTB

Visualization: EM-C; VCB; JE-L; MTB

Funding acquisition: AT; DK; AMV; TMC; MTB

Project administration: EM-C; AT; AMV; TMC; JE-L; MTB Supervision: EM-C; AT, OCF; AMV; TMC; JE-L; MTB

Writing – original draft: EM-C; VCB; CM; DK; JE-L; MTB

Writing – review &amp; editing: EM-C; VCB; CM; AT; OCF; RMP; DK; AMV; TMC; JE-L; MTB

## Competing interests

Authors declare that they have no competing interests

## STAR Methods

### Cohort and Study Design

All patients included in the present study sought testing at the Diagnostic Screening Center for COVID-19 at the Federal University of Rio de Janeiro (CTD-UFRJ) and declared written informed consent. From April to December 2020, we enrolled two thousand seven hundred and fifty-nine patients who were tested for SARS-CoV-2 infection at the Diagnostic Screening Center for COVID-19 of the Federal University of Rio de Janeiro (CTD-UFRJ). Among them, 1,133 individuals (41.07%) tested positive for the presence of SARS-CoV-2 RNA by quantitative PCR with reverse transcription (RT-qPCR) on nasopharyngeal swab samples. Those individuals were offered weekly follow-up testing until SARS-CoV-2 RNA was no longer detected. Blood from those patients was collected in heparinized tubes for plasma and PBMC storage and further analysis. Symptoms, use of medication, comorbidities and demographic information were assessed by oral questionnaire performed by trained personnel. Based on blood sample availability, 33 patients were selected from those with persistent SARS-CoV-2 infection, defined as positive SARS-CoV-2 RT-qPCR in upper respiratory tract (URT) samples after 21 days after symptom onset. As such, 32 patients were selected from those who did not display persistent viral RNA, defined as a negative SARS-CoV-2 RT-qPCR in URT samples up to 21 days after symptom onset. The criterion was guided by the median of positivity duration in the overall cohort, which was around three weeks (Voloch et al, 2021). Twenty-five non-infected volunteers were included as controls, defined as a negative SARS-CoV-2 qRT-PCR, no history of a positive SARS-CoV-2 qRT-PCR and no seroconversion for SARS-CoV-2 epitopes. All procedures and experiments were approved by the correspondent Ethic Committee Board (CAAE: 30161620.0.1001.5257; 4.245.490).

### PBMC and plasma isolation

PBMCs were isolated from blood collected in lithium heparin tubes by Ficoll–Hypaque density gradient centrifugation. Briefly, blood was layered on a density gradient (Hystopaque® 1077, Sigma-Aldrich), and PBMCs were separated by centrifuging at 400 rcf for 30 min. PBMCs were washed four times in phosphate-buffered saline (PBS), submitted to ACK for 5 min to lysate red blood cells, and frozen in liquid nitrogen in 90% fetal bovine serum (FBS) with 10% DMSO (Sigma-Aldrich) until used for flow cytometry and stimulation assays. Plasma samples were collected in 10-ml tubes in lithium heparin tubes, centrifuged at 400 rcf for 10 min, aliquoted, and stored at −20°C for further experiments.

### Cell culture

PBMCs were thawed in RPMI 1640 medium (Lonza) supplemented with 10% FBS (Gibco™) and penicillin (100 IU/ml; Gibco™)/streptomycin (100 μg/ml; Gibco™). For T cell stimulation, 0.5×10^6^ PBMCs were cultured in 500µl of medium in a 24-well flat-bottomed microplate and stimulated for 4 days with anti-CD3/anti-CD28 Dynabeads™ Human T-activator (10 μL/mL, Gibco™). For detection of SARS-CoV-2–specific CD4^+^ and CD8^+^ T cells, 0.5×10^6^ PBMC cells were stimulated in a 96-well U-bottom plate for 7 days in 200 μl of medium containing 1 μg/ml of SARS-CoV-2 variant Spike peptide pools (JPT Peptide Technologies) in the presence of IL-2r (20UI, PrepoTech Inc.). As a control, some cells were maintained with IL-2r alone. After 6 days, the cells were re-stimulated with 10μg/ml of the aforementioned peptide pools overnight. In order to optimize the detection of intracellular cytokines, Brefeldin A (10 μg/mL; BD Biosciences, San Diego, CA, USA) was added in the last 4 h (for Dynabeads™) or 12h (for SARS-CoV-2 peptide pools) of the cell cultures. Cultures were maintained in a humidified incubator with 5% CO2 at 37°C. After stimulation, cells were stained for proliferation and/or phenotypic lymphocyte markers by Flow Cytometry.

### Flow Cytometry

Staining for flow cytometry analysis was performed using fluorescently-labeled specific anti-human antibodies. Unless otherwise stated, antibodies were purchased from Biolegend: IgD-PE-Cy7, CD15-BV421, CD56-PE-CF594, CD14-PECy5, CD16-BV786, HLA-DR-BB515/PECy5 (BD Bioscience), CD11b-APC-Cy7, CD304-PE, CD11c-BV711, CD1c-APC, CD141-BV650, CD3-BV510/BV605, CD4-BV785, CD19-BV421/BV510, CD8-BV711/APC-Fire750, CD27-FITC/PE, CCR7-BV421, CD45RA-PE-Cy7/PE-Cy5, CD25-AF700, PD-1-PE, CD38-BV711/PE-Cy5, CXCR5-PE-CF594, Ki67-APC, IL-10-BV421, Granzyme B-FITC (BD Bioscience), IFNγ-APC-Fire750, TNFα-BV650, IL-9-PE, IL-6-PE/PE-Cy7, and IL-21-PerCP-Cy5.5.

Briefly, 5×10^5^ cells were incubated with LIVE/DEAD Fixable Aqua Dead Cell staining (1:1000, BV510; Invitrogen) in PBS for 30 min, and washed with PBS containing 3% FBS and 0.01% sodium azide. Next, cells were incubated with the fluorescently-labelled antibodies for 30 min at room temperature in the dark. When intracellular staining was required, cells were permeabilized with the Cytofix/Cytoperm solution (BD Pharmigen) at 4°C for 20 min and subsequently incubated for 30 min at 4°C with the appropriate antibodies. Events were acquired on LSR FORTESSA X-20 (BD Biosciences). Target cells were gated based on forward and side scatter properties, singlets and living cells were analyzed by using FlowJo^©^ Software. FMO controls and single-stained samples were used to periodically check the settings and gates on the flow cytometer.

### Quantification of inflammatory, anti-inflammatory, and antiviral mediators

Cytokines and immune mediators were quantified with a multiplex magnetic bead-based immunoassay according to the manufacturer’s instructions, using the Bio-Plex Pro™ Human Cytokines 27-Plex Assay (BIO-RAD) and the Human ProcartaPlexTM kit (Invitrogen™, ThermoFisher Scientific), including the following analytes: basic FGF, Eotaxin, G-CSF, GM-CSF, IFN-γ, IL-1β, IL-1RA, IL-2, IL-4, IL-5, IL-7, IL-8, IL-9, IL-10, IL-12 (p70), IL-13, IL-15, IL-17A, IP-10/CXCL10, MCP-1/CCL2, MIP-1α/CCL3, MIP-1β/CCL4, PDGF-BB, TNF-α, and VEGF, IFNα, IFNβ, IFNΩ, IL-29, IL-18, IL-21, syndecan, ferritin. Data were acquired in a Luminex MAGIPIX® System (serial number MAGPX11266003) using the XPONENT software. Additionally, IL-6, and RANTES/CCL5 concentrations were determined in plasma samples by enzyme-linked immunosorbent assay (Human IL-6 and RANTES/CCL5 TMB ELISA, Peprotech), according to the manufacturer’s instructions. Absorbance was measured in a SpectraMax^®^ microplate reader (Molecular Device).

### Quantitative PCR

RNA from PBMCs was extracted using Fastzol (QuatroG, Brazil), according to the manufacturer’s instructions. 1µg of RNA was reverse transcribed to complementary DNA (cDNA) with the High-Capacity RNA-to-cDNA™ Kit (Applied Biosystems™) and random primers. Quantitative RT-PCR was performed with the Power SYBR^®^ Green PCR Master Mix (Applied Biosystems™) in an ABI 7500 instrument (Applied Biosystems™). Ct values were normalized to the mRNA expression of GAPDH and relative expression was calculated with the ΔΔCt method. Primer sequences are available upon request.

### Determination of SARS-CoV-2 specific immunoglobulins and neutralizing antibodies

Anti-SARS-CoV-2 spike protein IgM, IgA, and IgG antibodies were quantified by enzyme-linked immunosorbent assay, following the *S-UFRJ test* protocol, as previously described ^48^. Briefly, serial dilutions of plasma samples (starting 1:40, in PBS 1% BSA) were incubated in high-binding ELISA plates previously coated with 50 μL of SARS-CoV-2 spike protein (4 μg/mL) overnight. Antibodies were detected with goat anti-human IgG, IgA, and IgM (Fc)-horseradish peroxidase antibodies (Southern Biotech) and developed with TMB (Scienco). Optical density (OD) was measured in a SpectraMax® microplate reader (Molecular Device, USA). Plasma titers of neutralizing antibodies were quantified with a multiplex magnetic bead-based immunoassay, using the Bio-Plex Pro Human SARS-CoV-2 Neutralization Antibody Assay (BIO-RAD) according to the manufacturer’s instructions.

### Statistical analysis

One-dimensional probability distributions of samples were analyzed by Kolmogorov– Smirnov test. Statistical analysis was performed by one-way variance analysis followed by Bonferroni post-test for samples with normal distribution; or Kruskal-Wallis analysis followed by Dunn post-test. Correlations were analyzed by Spearman’s rank correlation coefficient. The statistical analysis was performed using Prism 8.0 software (GraphPad Software, San Diego, CA).

## Supplementary Material

**supplementary Fig. 1.**
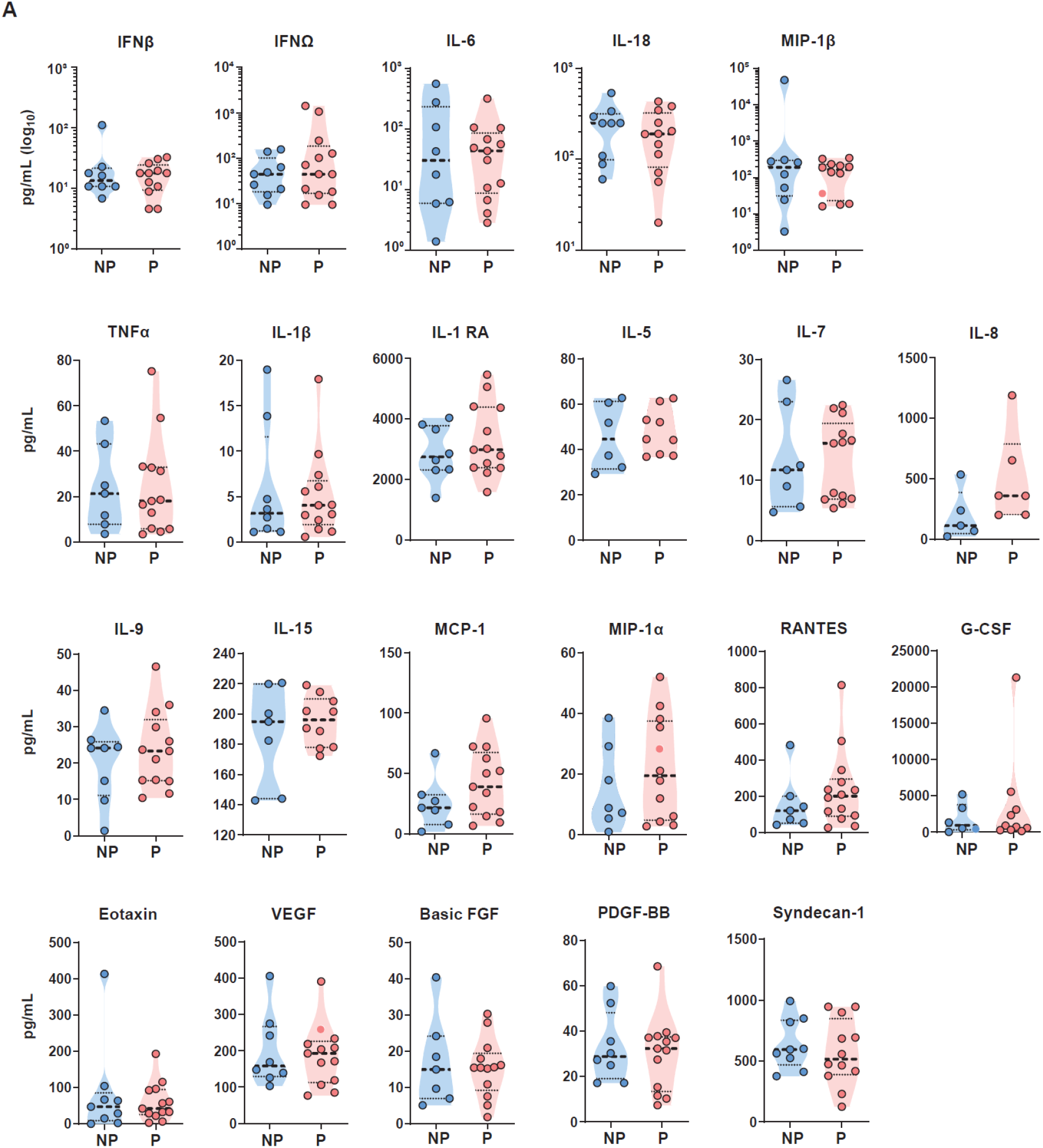
Cytokine analysis in swab samples at disease onset. Cytokine quantification in swabs samples from upper respiratory tract of NP (n=9) and P (n=13) at <10 DSSO by multiplex magnetics bead-based immunoassay. Each dot represents a subject. Filled dots represent a positive qRT-PCR of nasopharyngeal samples for SARS-CoV-2. Statistical significance was calculated using Mann-Whitney test.

**supplementary Fig. 2.**
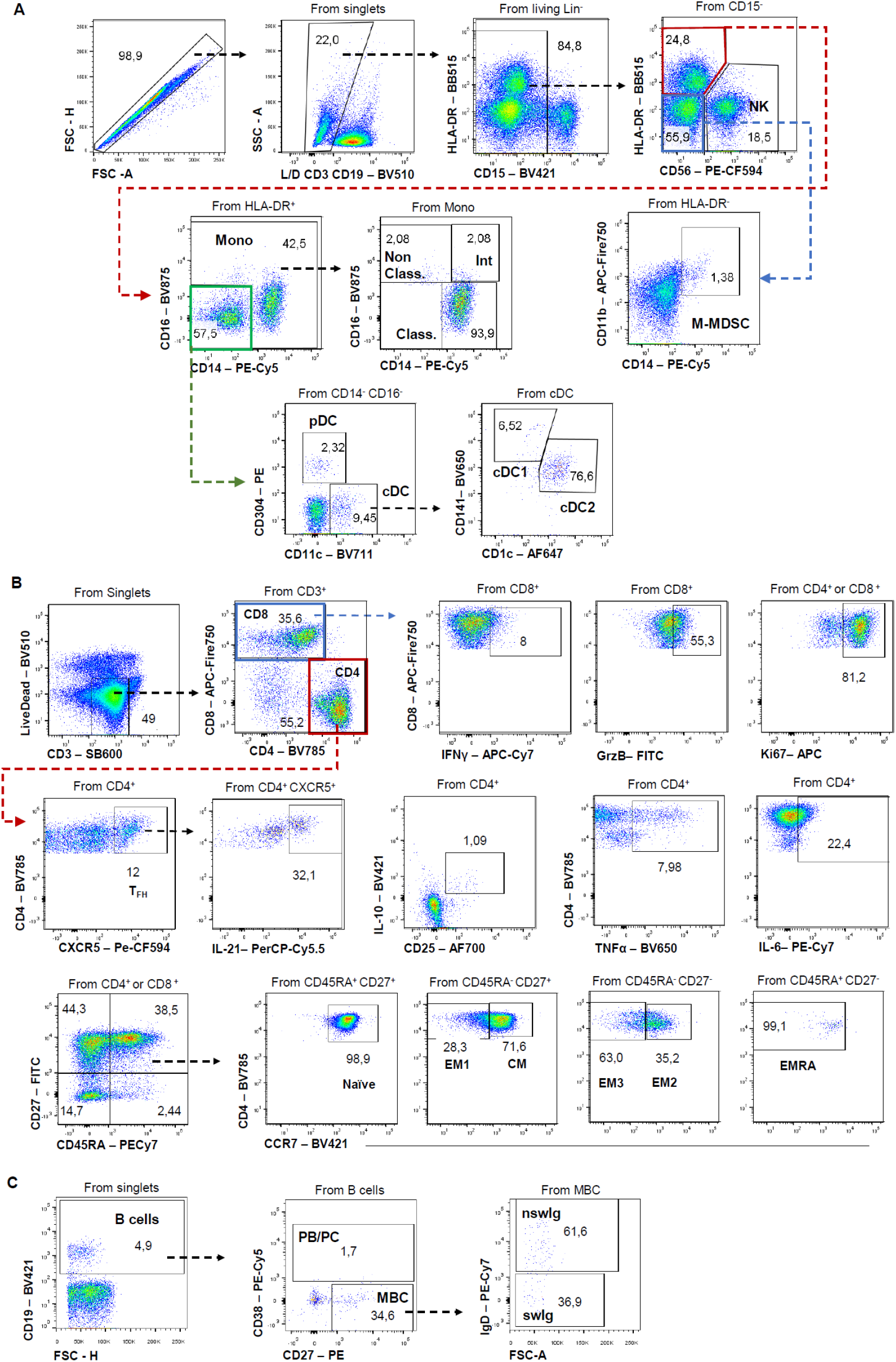
Flow cytometry gating strategies. Gating strategies on PBMCs for immunophenotyping of (A) innate immune cells, among them NK cells, monocytes and dendritic cells, (B) T lymphocytes and (C) B lymphocytes.

**supplementary Fig. 3.**
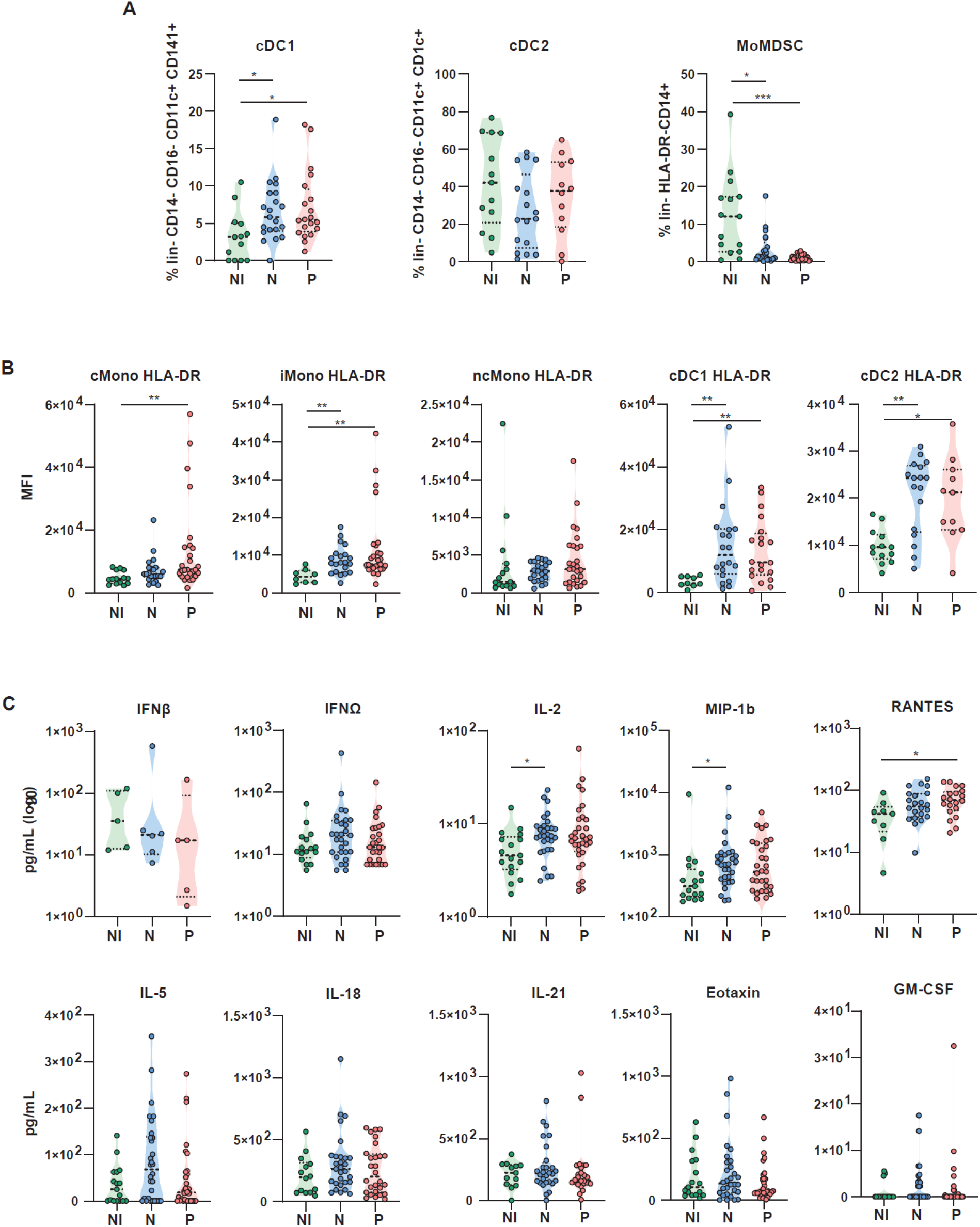
Immunophenotyping and plasma cytokine analysis at disease onset. Immunophenotyping of major innate immune cell populations in PBMC (A) LIVE/DEAD^-^CD3^-^ CD19^-^CD56^-^ (lin^-^) CD14^-^CD16^-^CD11c^+^CD141^+^ type 1 dendritic cells (cDC1), LIVE/DEAD^-^lin^-^ CD14^-^CD16^-^CD11c^+^CD1c^+^ type 2 dendritic cells (cDC2) and LIVE/DEAD^-^lin^-^HLA^-^DR^-^CD14^+^ myeloid-derived suppressor cells (MoMDSCs). (B) MFI of HLA-DR in cMono, iMono, ncMono, cDC1 and cDC2 cell populations. (C) Quantification of plasma cytokines in non-infected individuals (NI), non-persistent patients (NP) and persistent patients (P) at <10 DSSO by multiplex magnetics bead-based immunoassay. Each dot represents a different subject. Statistical significance was calculated using Mann-Whitney test or Kruskal-Wallis analysis followed by Dunn post-test and indicated by *p % 0.05; **p % 0.01; and ***p % 0.001. DSSO, Days since symptom onset.

**supplementary Fig. 4.**
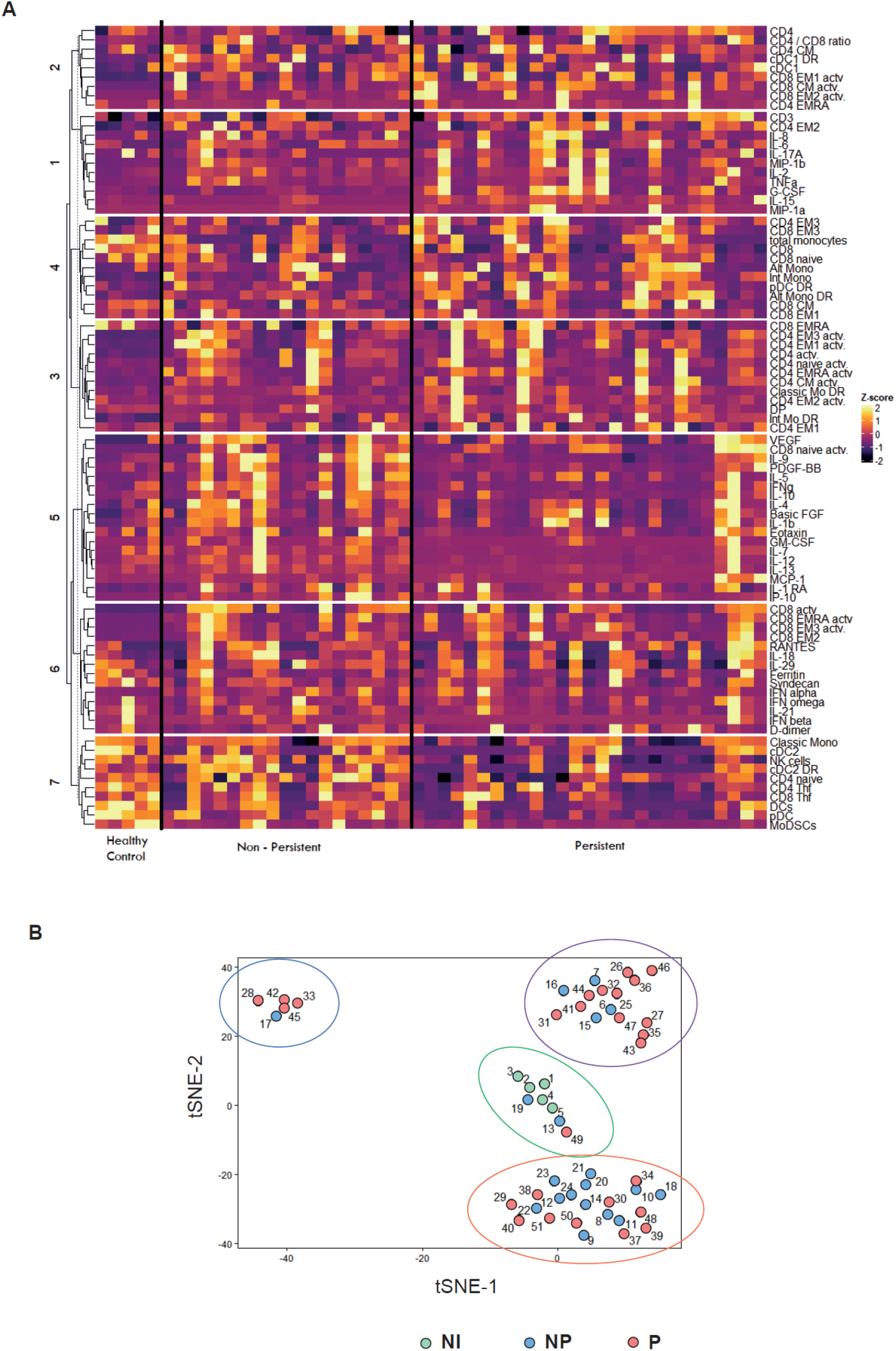
Cluster analysis of immunophenotyping and plasma cytokine data at disease onset. (A) Heat map of blood cell populations and cytokine concentration in serum from P, NP and NI at <10 DSSO measured by flow cytometry and Luminex assay. K-means clustering was used to determine cytokine clusters 1-7). Measurements were normalized across all patients by Z-score. (B) t-Distributed Stochastic Neighbor Embedding (t-SNE) probabilistic dimensionality reduction technique applied in PBMC immunophenotyping and cytokines concentration at <10 DSSO from 51 patients (P, Blue, *n*=27; NP, Orange, *n*=19; NI, Green, *n*=5). Measurements were normalized across all patients by Z-score.

**supplementary Fig. 5.**
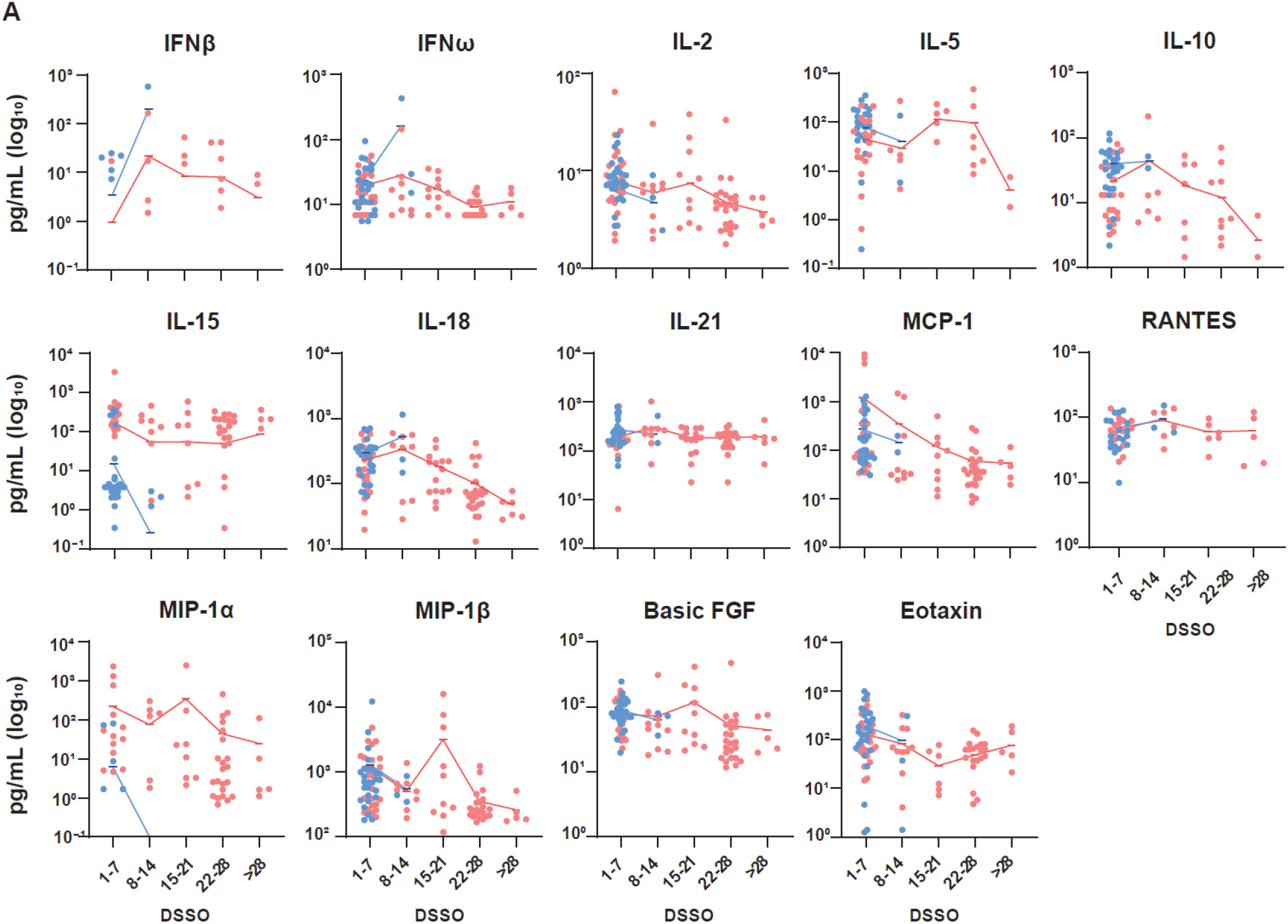
Longitudinal cytokine analysis. (A) Weekly longitudinal quantification of plasma cytokines in non-persistent (NP, blue) and persistent (P, red) patients by multiplex magnetics bead-based immunoassay.

**supplementary Fig. 6.**
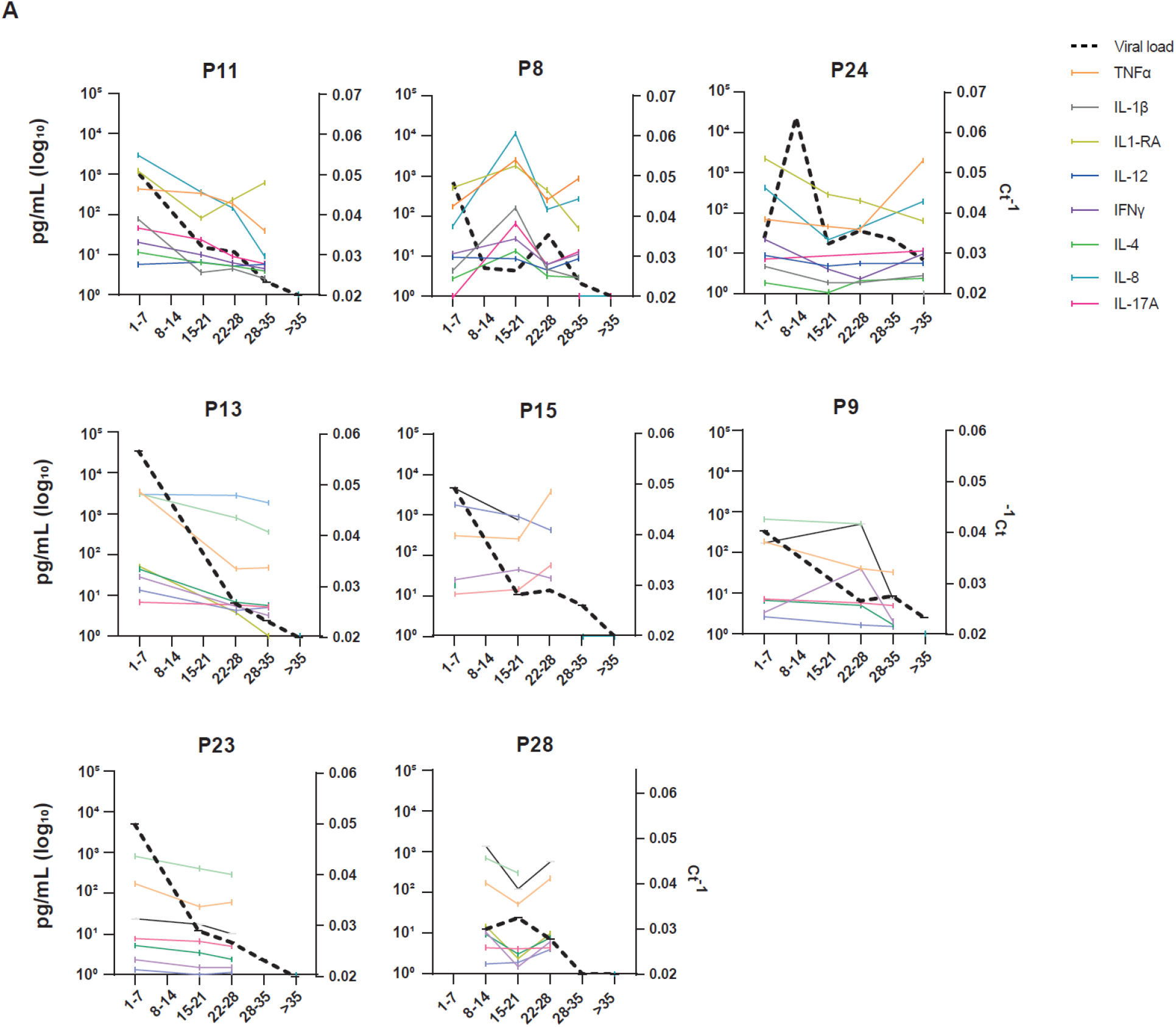
Longitudinal analysis of plasma cytokine and viral loads in individual patients. (A) Longitudinal depiction of plasma IFNα, IP-10, TNFα, IL-1β, IL-12, IFNγ, IL-4, IL-9, IL-17A and IL-8 and viral loads in individual patients with persistent infection and at least three longitudinal samples available. Ct values (depicted as Ct^-1^) were determined by qRT-PCR of viral RNA on nasopharyngeal swab samples. Cytokine concentrations were measured by multiplex magnetics bead-based immunoassay.

**supplementary Fig. 7.**
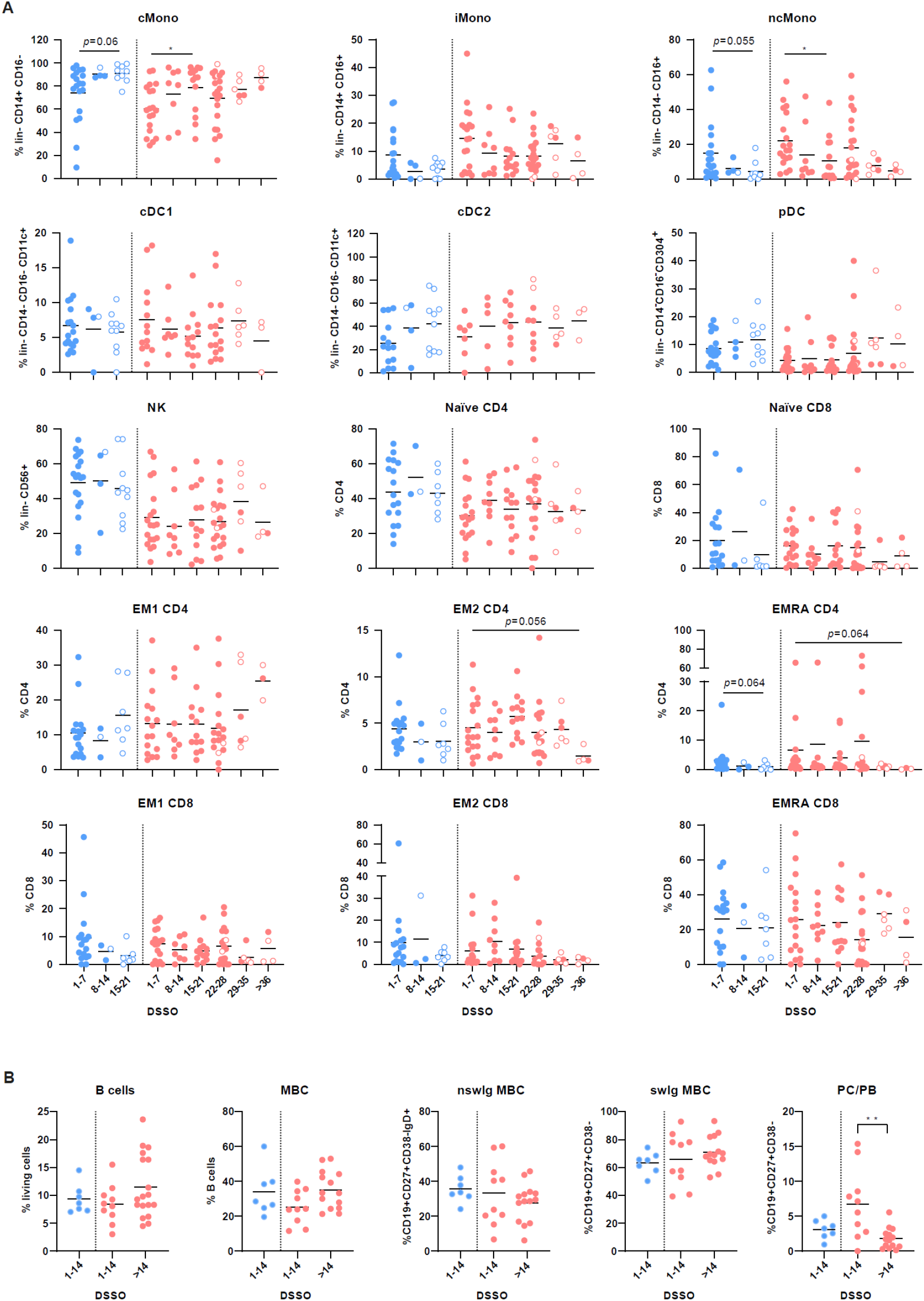
Longitudinal analysis of immune cell populations. Longitudinal immunophenotyping in PBMC of NP and P patients. Filled dots represent individual samples longitudinally collected at different time points until resolution of infection from patients positive by qRT-PCR for SARS-CoV-2, while empty dots represent samples from convalescent patients coinciding with the first negative qRT-PCR for SARS-CoV-2. Statistical significance was calculated using Mann-Whitney test or Kruskal-Wallis analysis followed by Dunn post-test and indicated by *p % 0.05; **p % 0.01; and ***p % 0.001.

**supplementary Fig. 8.**
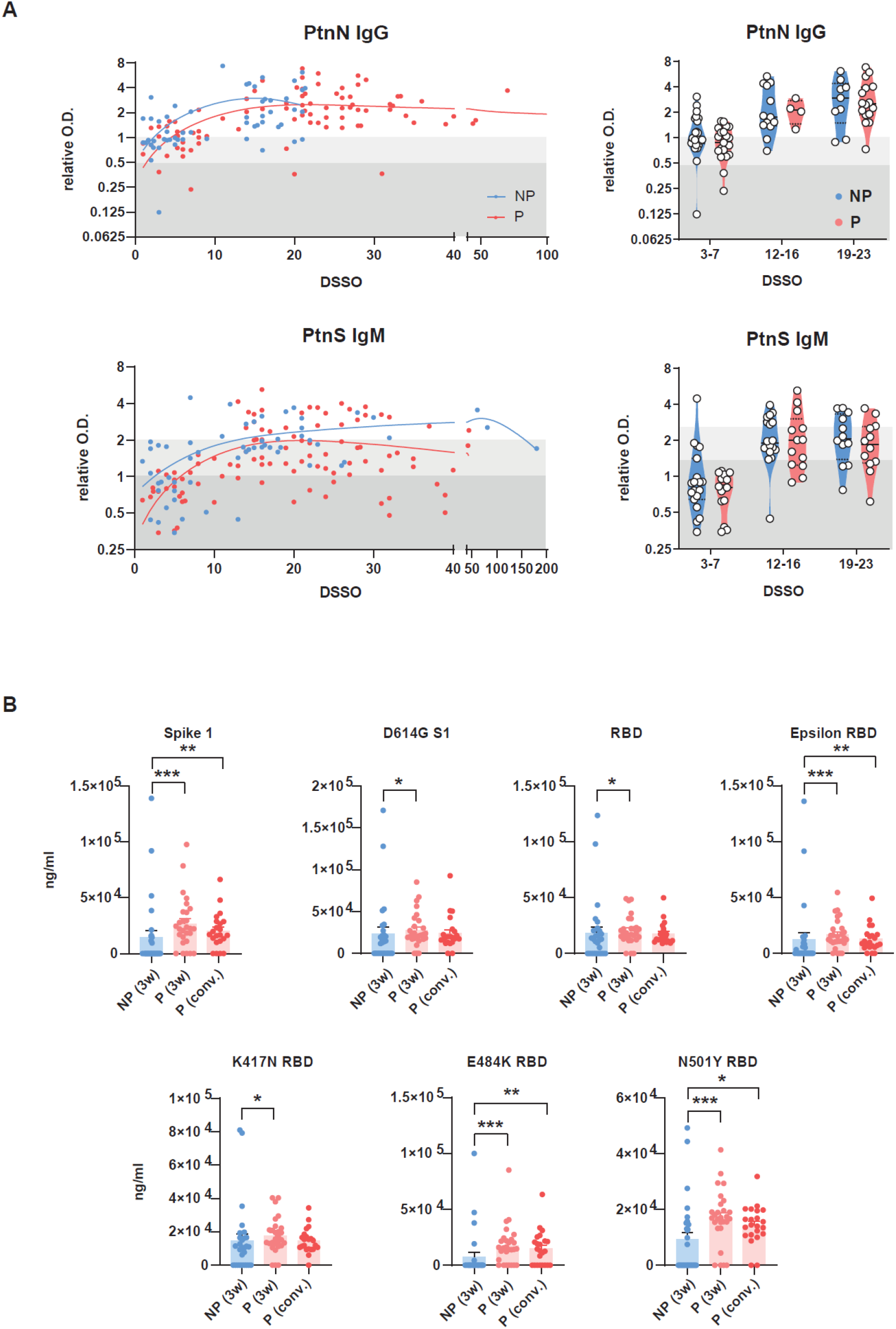
Longitudinal analysis of SARS-CoV-2-specific immunoglobulins and neutralizing antibodies. Longitudinal assessment of anti-N IgG and anti-S IgM antibodies in plasma from P and NP by ELISA assay. (H) Titers of neutralizing antibodies against S1, D614G S1, RBD, Epsilon RBD, K417N RBD, E484K RBD, and N501Y RBD variants of SARS-CoV-2 in plasma from NP and P around 21 DSSO and P by the time of viral clearance by competitive Luminex multiplex neutralization assay. Statistical significance was calculated using Mann-Whitney test or Kruskal-Wallis analysis followed by Dunn post-test and indicated by *p % 0.05; **p % 0.01; and ***p % 0.001. DSSO, Days since symptom onset.

## Notes

### Competing Interest Statement

The authors have declared no competing interest.

### Funding Statement

This study was funded by ANRS | Maladies infectieuses emergentes/INSERM grant (MUCOVID-007) (to DK and MTB); by CAPES, Edital de Selecao Emergencial II CAPES grant 88887.507381/2020-00 (to MTB); by Fundacao de Amparo a Pesquisa do Estado do Rio de Janeiro (FAPERJ), grant E-26/201.128/2022 (272688) and E-26/211.564/2019 (252360) (to MTB), E-26/010.002434/2019 and E-26/210.178/2020 (to AT), E-26/203.002/208 (to A.M.V.); by Conselho Nacional de Desenvolvimento Cientifico e Tecnologico (CNPq) grant 312477/2021-0 (to MTB) and grants 439649/2018-8 and 316796/2021-2 (to A.M.V.); Instituto Serrapilheira (to AT).

### Author Declarations

CONEP (National Ethics Committee for Research) and is entitled Characterization of risk factors and development of new serum tests for SARS-CoV-Infection Ethic Committee Board (CAAE: 30161620.0.1001.5257; 4.245.490)

